# Immune response to COVID-19 vaccination is attenuated by poor disease control and antimyeloma therapy with vaccine driven divergent T cell response

**DOI:** 10.1101/2021.10.21.21265158

**Authors:** Karthik Ramasamy, Ross Sadler, Sally Jeans, Paul Weeden, Sherin Varghese, Alison Turner, Jemma Larham, Nathanael Gray, Oluremi Carty, Joe Barrett, Stella Bowcock, Udo Oppermann, Gordon Cook, Chara Kyriakou, Mark Drayson, Supratik Basu, Sally Moore, Sarah McDonald, Sarah Gooding, Muhammad K Javaid

## Abstract

**Background:** Myeloma patients frequently respond poorly to bacterial and viral vaccination. A few studies have reported poor humoral immune responses in myeloma patients to COVID-19 vaccination.

**Methods:** Using a prospective study of myeloma patients in UK Rudy Study cohort, we assessed humoral and Interferon gamma release assay (IGRA) cellular immune responses to COVID-19 vaccination post second COVID-19 vaccine administration.

**Findings:** We report data from 214 adults with myeloma (n=204) or smouldering myeloma (n=10) who provided blood samples at least 3 weeks after second vaccine dose. Positive Anti-Spike antibody levels (> 50 IU/ml) were detected in 189/203 (92.7%), positive IGRA responses were seen in 97/158 (61.4%) myeloma patients. Only 10/158 (6.3%) patients were identified to have both a negative IGRA and negative Anti-Spike protein antibody response. 95/158 (60.1%) patients produced positive results for both anti-Spike protein serology and IGRA. After adjusting for disease severity and myeloma therapy, poor humoral immune response was predicted by male gender. Predictors of poor IGRA included anti-CD38/ anti-BCMA therapy and Pfizer-BioNTech (PB) vaccination.

**Interpretation:** Significant majority of myeloma patients elicit Anti-Spike protein antibody responses to COVID-19 vaccine with 60% of myeloma patients showing both humoral and T cell response. Predictors of a poor immune response included male gender, myeloma therapy regimen and administration of Pfizer-BioNTech vaccination. Further work is required to understand the clinical significance of divergent cellular response to vaccination.

**Funding:** Funding for this study has been received from Blood Cancer Vaccine Consortium and Janssen UK. RUDY platform has been funded by NIHR.

## Introduction

High mortality rates with COVID-19 in myeloma patients coupled with anticipated poor COVID-19 vaccine responses will increase isolation periods and be detrimental to their myeloma care^1^. Myeloma patients have been shielding since the start of the COVID-19 pandemic. This has resulted in disruption to therapy and healthcare provision, significant social isolation and worse mental health^2,3^. The solution to this is providing meaningful protection against COVID-19 through vaccination and ensuring this protection is durable. If vaccination does not offer protection, then what alternative pharmacological strategies in addition to behavioural practices to keep myeloma patients safe, must be explored.

Myeloma is a malignant clonal proliferation of post germinal centre B cells and plasma cells. The canonical role of these immune cells is to provide both immunity against pathogens, respond to vaccination and deal with pathogens in mucosal surfaces with IgA production. Prolonged SARS-Cov2 viral shedding and delayed or negligible seroconversion has been observed in infected patients^4^. Myeloma commonly affects the elderly population, a cohort that principally suffer with co-morbidities and immunosenescence, further increasing their risk of developing complications from infections and poor response to vaccination. Myeloma as a disease requires ongoing immune suppressive chemotherapy to induce and maintain remission which results in frequent medical visits and exposure to hospital environment. Influenza vaccination studies report that myeloma patients often require repeated doses of vaccine to mount optimal antibody response ^5^. Pneumococcal vaccination response in myeloma is frequently impaired and was found to be associated with poor disease control in a study of myeloma patients given 23-valent polysaccharide vaccine ^6^. The poor seroconversion that results from established cellular and humoral immune dysfunction can be exacerbated by active treatment and disease status ^7^. Humoral immune response to COVID-19 vaccination in myeloma patients given 2 doses of mRNA based COVID-19 vaccination ranged between 80 – 85% in 3 separate cohort studies ^8,9,10^. However, the protective titre of antibodies required to prevent re-infection (neutralisation) is unclear, as is the ability to protect patients from other SARS-COV2 virus variants of concern (VOC). Moreover, the role of the cellular immune response to SARS-COV2 is crucial, and to date has not been assessed in a large cohort of MM patients. Robust T cell responses could enhance durability of humoral responses, which is important in an ongoing pandemic generating new VOC.

In the UK both mRNA based (Pfizer Biontech; PB) and viral vector-based vaccination (Oxford/ Astra Zeneca (AZ)) were offered to patients. Also, the recommended schedule of 12 weeks for second dose was divergent from manufacturer’s recommendations. It is unclear whether these play a role in vaccination responses in myeloma patients. Understanding drivers of poor response to vaccination and potential salvage strategies (i.e., booster, passive antibody) for this subset is key to managing their myeloma optimally. As the vast majority of patients with myeloma require ongoing therapy; the effect of both myeloma response and myeloma therapy on both eliciting and durability of vaccine responses requires prospective evaluation.

In order to address these evidence gaps, all of which directly affect patient management decisions, both immediately and booster dose planning, we initiated a national web-based prospective study of adults with MM to determine the humoral and cellular responses post the completion of the first and second dose of COVID-19 vaccine schedule administered in the UK in 2021.

## Methods

This is a prospective, observational study. The study is based on the existing RUDYstudy.org platform (LREC 14/SC/0126 & RUDY LREC 17/SC/0501), an established online rare disease platform with online dynamic consent and patient reported data^11^.

### Recruitment

To ensure reaching our recruitment target rapidly, multiple pathways for recruitment were employed. The study is currently open to any UK resident to join via 30 hospitals and through Myeloma UK national patient charity and Blood Cancer UK digital platform. At the hospitals, the clinical team were able to include the letter of invitation and study flyer with hospital appointment letters, and then offer consent to participants during that clinic visit or direct them to the study website for online registration and consent. Participants were requested to identify their lead hospital clinician. Where the lead clinician was a known haematologist, participants were considered to have a confirmed MM diagnosis. Informed online dynamic consent was obtained for all participants.

### Assessment

Participants were able to enter online the time of their first and second vaccine doses. Based on these dates, a sample box was posted to the participants, containing serum, EDTA and heparin blood tubes after the second vaccine dosing. Participants took the sample box to have their bloods taken either at their primary care or secondary care clinic. Primary care clinicians were reimbursed for their time in taking part in the study. Samples were centrally received, logged, processed and either distributed to collaborators for assays, or appropriately stored at the Immunology Lab, Churchill Hospital, Oxford (Figure 1).

**Figure 1.**
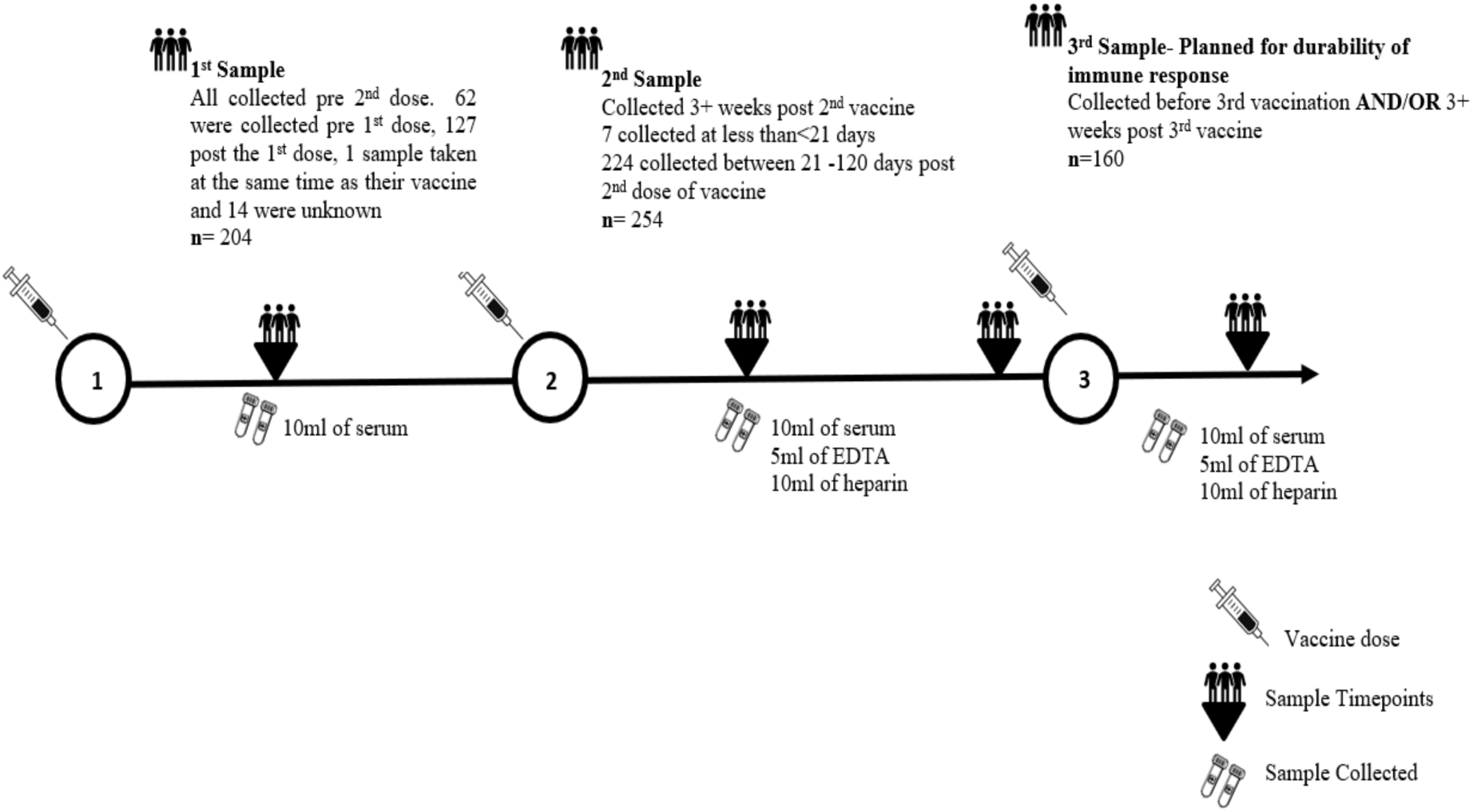
Blood Sampling plan aligned to vaccination with sample numbers and type of samples obtained.

In addition, participants completed online questionnaires that included COVID symptoms, testing and myeloma features such as current myeloma status (see supplementary materials). Independently, there was review of clinical data to identify myeloma status and treatment at the time of second vaccine dose in a subset of participants. Myeloma status was coded as Complete remission/ Very good partial remission (CR/VGPR); Partial remission/ Stable disease (PR/Stable); and Progression/ Relapse. Chemotherapy treatment was coded as proteasome inhibitors - ixazomib, carfilzomib, bortezomib; immunomodulatory drugs - thalidomide, lenalidomide, pomalidomide; anti-CD38 antibody (daratumumab, Isatuximab); and anti-BCMA targeted treatment – (belantamab), Others - bendamustine, cyclophosphamide, dexamethasone, other steroids. If the clinical data were not available for myeloma status, the closest patient reported myeloma status was used.

### Laboratory assessments

Collected serum samples were tested for antibodies against SARS-CoV2 nucleocapsid (N) or spike (S) protein. SARS-CoV2 N protein antibodies were measured by turbidimetry (Abbott), with samples that produced values of >1.4IU/ml considered to be positive. SARS-CoV2 S protein antibodies were measured by turbidimetry (Abbott) (IgG serology only), with a cut-off value of 50IU/ml considered to be a positive result. Heparin samples collected from patients were used to isolate peripheral blood mononuclear cells (PBMCs), which were then placed into a SARS CoV2 S protein interferon gamma release assay (IGRA) (Oxford immunotec T IGRA) to quantify SARS-CoV2-specific effector T cells. The assay was followed as per the kit insert with positive results defined as >8 interferon gamma releasing cells/ 10^6^ PBMCs.

### Statistical analysis

Descriptive statistics included t-tests and ANOVA for parametric outcome measures, Spearman correlations, and Kruskall-Wallis tests for non-parametric outcomes. Categorical results were evaluated by Chi-Squared and Fisher’s exact tests when individual cell counts were less than 10. We tested the agreement of clinician reported myeloma status with the closest participant self-reported myeloma status and for patient reports within 90 days of the second vaccine dose. Multivariate logistic regression analyses were performed to identify independent predictors of Anti-S antibody response and IGRA positivity. We combined humoral and T cell outcomes responses to generate 4 independent groups: combined positive Anti-S antibody and IGRA reactivity compared with those with either Anti-S antibody or IGRA reactivity and those with double negative results. These groups were analysed with multinomial regression. Significance was determined as p< 0.05.

## Results

Two hundred and fourteen participants with myeloma (n=204) or smouldering myeloma (n=10) completed the COVID-19 questionnaire and returned a blood sample at least 3 weeks after their second dose of COVID-19 vaccine (median 9.5 weeks (range 3 – 20.4 weeks). The baseline characteristics are shown in Table 1, with over 30% of participants aged 70 years and over. The type of vaccine used was reported by 160 participants with the AZ vaccine reported by 59.6% of patients with myeloma and 44.4% of patients with smouldering myeloma and the PB vaccine used by the remainder. Neither age, sex, myeloma status or chemotherapy at time of vaccination predicted the type of vaccine used (p>0.1). However, the PB vaccine was given an average of 6 days before AZ vaccine (p=0.01).

**Table 1.**
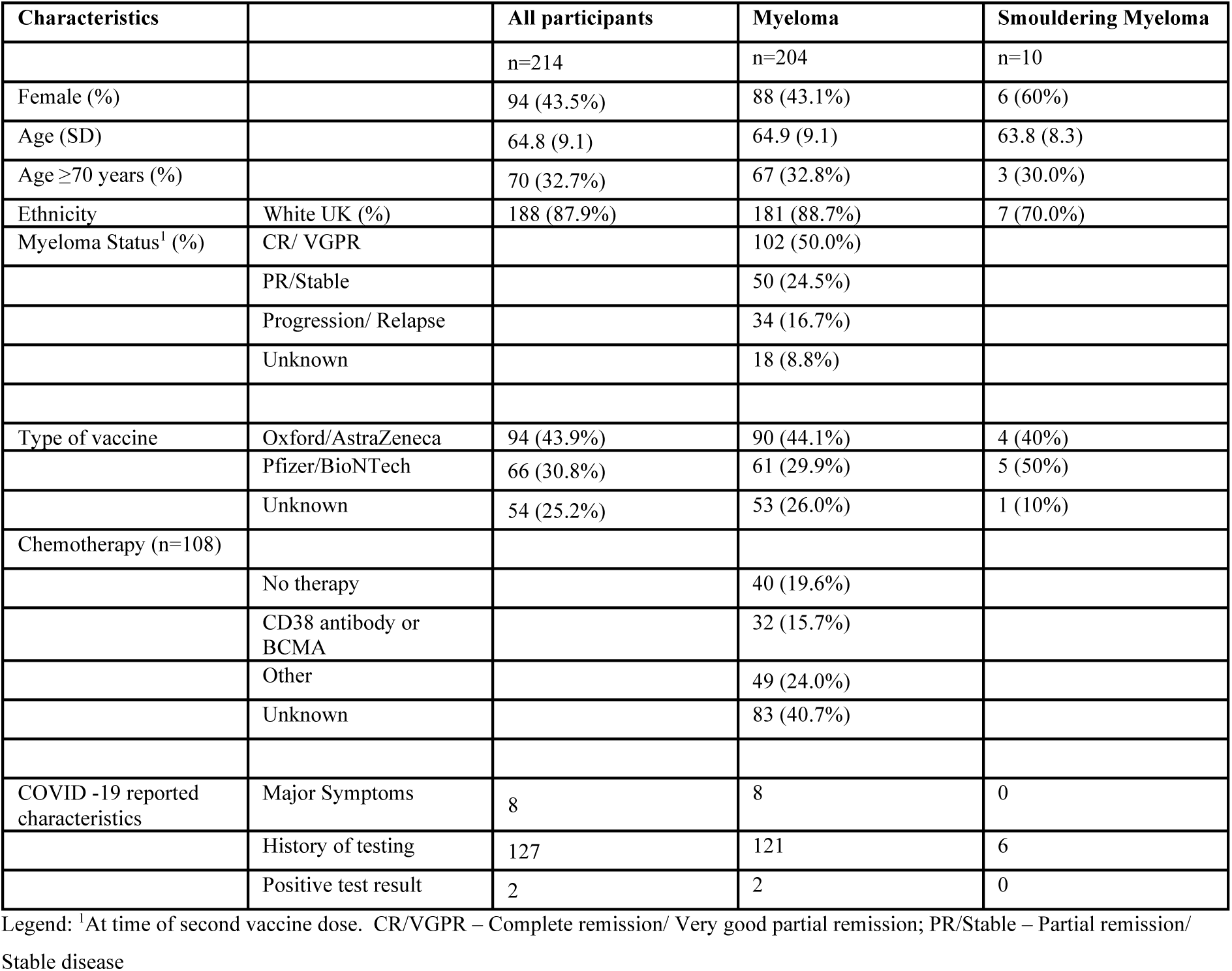
Baseline characteristics of patients included in the analysis of immune response post vaccination.

| Characteristics |  | All participants | Myeloma | Smouldering Myeloma |
| --- | --- | --- | --- | --- |
|  |  | n=214 | n=204 | n=10 |
| Female (%) |  | 94 (43.5%) | 88 (43.1%) | 6 (60%) |
| Age (SD) |  | 64.8 (9.1) | 64.9 (9.1) | 63.8 (8.3) |
| Age ≥70 years (%) |  | 70 (32.7%) | 67 (32.8%) | 3 (30.0%) |
| Ethnicity | White UK (%) | 188 (87.9%) | 181 (88.7%) | 7 (70.0%) |
| Myeloma Status<sup>1</sup> (%) | CR/ VGPR |  | 102 (50.0%) |  |
|  | PR/Stable |  | 50 (24.5%) |  |
|  | Progression/ Relapse |  | 34 (16.7%) |  |
|  | Unknown |  | 18 (8.8%) |  |
| Type of vaccine | Oxford/AstraZeneca | 94 (43.9%) | 90 (44.1%) | 4 (40%) |
|  | Pfizer/BioNTech | 66 (30.8%) | 61 (29.9%) | 5 (50%) |
|  | Unknown | 54 (25.2%) | 53 (26.0%) | 1 (10%) |
| Chemotherapy (n=108) |  |  |  |  |
|  | No therapy |  | 40 (19.6%) |  |
|  | CD38 antibody or BCMA |  | 32 (15.7%) |  |
|  | Other |  | 49 (24.0%) |  |
|  | Unknown |  | 83 (40.7%) |  |
| COVID -19 reported characteristics | Major Symptoms | 8 | 8 | 0 |
|  | History of testing | 127 | 121 | 6 |
|  | Positive test result | 2 | 2 | 0 |

Legend: <sup>1</sup>At time of second vaccine dose. CR/VGPR – Complete remission/ Very good partial remission; PR/Stable – Partial remission/ Stable disease

The humoral response findings after the first and second COVID-19 vaccine dose are shown in Table 2, with the distribution of individual antibody results for all participants shown in Figure 2. Serological evidence of prior COVID infection, by the detection of anti-N antibodies, after the second vaccine dose was found in 7 participants, 6 with a diagnosis of myeloma and 1 with a diagnosis of smouldering myeloma. No individual acquired Anti-N positive status between the first and second vaccine dose. Those with a positive Anti-N antibody (natural infection) at second sample had a significantly higher Anti-S protein response (p=0.002) (Supplementary figure 1).

**Table 2.** COVID-19 S and N-Protein Antibody status after first and second vaccine dosing.

|  |  | All participants | Myeloma | Smouldering Myeloma |
| --- | --- | --- | --- | --- |
| Post 1<sup>st</sup> vaccine dose |  | n=154 | n=146 | n=8 |
|  | S-protein antibody ≥50 IU/ml* | 50 (67.6%)* | 46 (66.7%)* | 4 (80%)* |
|  | N- protein antibody ≥ 1.5 IU/ml | 5 (3.3%) | 5 (3.4%) | 0 |
| Post 2<sup>nd</sup> vaccine dose |  | n=214 | n=203 | n=10 |
|  | S-protein antibody ≥50 IU/ml | 198 (92.5%) | 189 (92.7%) | 9 (90.0%) |
|  | N- protein antibody ≥1.5 IU/ml | 7 (3.3%) | 6 (3.0%) | 1 (9.1%) |

Legend: \*Restricted to 73 participants (69 myeloma and 4 smouldering myeloma) whose sample was at least 21 days after the 1<sup>st</sup> dose

**Figure 2.**
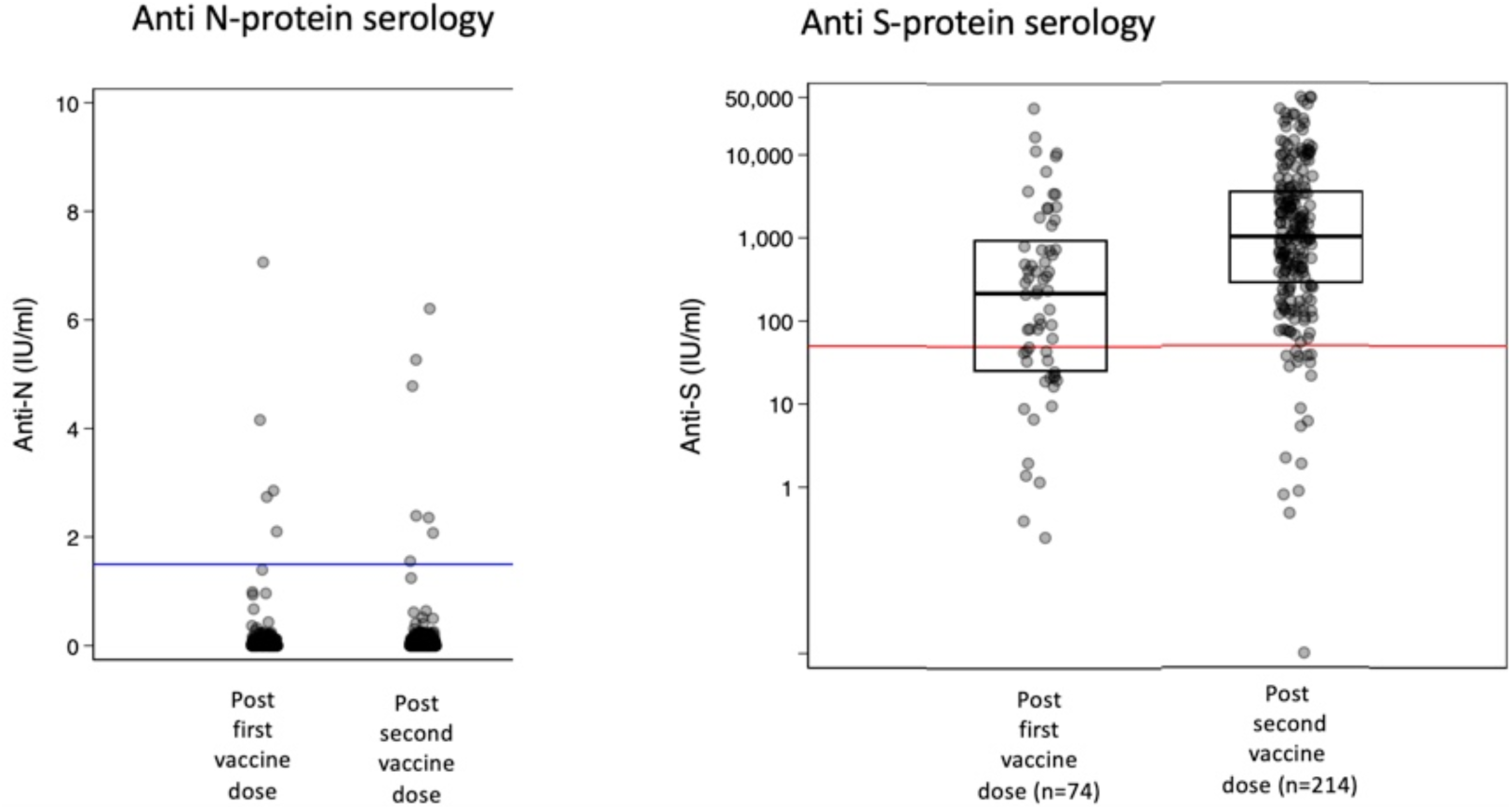
Post vaccination S and N protein antibody levels.

The comparison between S-protein antibody levels after the first and second vaccine are shown in Figure 3. One participant had sufficient Anti-S antibody after the first vaccine dose (Anti-S 1,268 IU/ml) but not after the second dose (Anti-S 41.2 IU/ml). This patient had smouldering myeloma and received a dose of rituximab for an inflammatory arthritis between vaccine doses. Anti-S levels by myeloma stage and in smouldering myeloma are shown in Figure 4. A low Anti-S concentration was more common in patients with partial response/stable disease or progressive/ relapse compared with those in complete remission/ very good partial remission (11.9% vs 3.9%, p=0.051). Men with myeloma were significantly more likely to have low Anti-S levels (<50 IU/ml) after second vaccination compared with women (11.3% vs 2.3%, p=0.015), with no difference by age (p=0.46). Anti-S levels were overall negatively associated with age (Supplementary figure 2a). Participants who reported receiving the PB vs AZ vaccine were no different in achieving a satisfactory Anti-S concentration (89.4% vs 93.6% respectively) but had higher Anti-S antibody concentrations (p=0.018) (Supplementary figure 2b). The time difference between the second vaccine dose and sample collection date was negatively correlated with Anti-S concentration (Spearman rho= -0.21, p=0.002) but not with Anti-S concentrations above or below 50 IU/ml (p=0.78) (Supplementary figure 3a). The median interval between the 1st and 2nd vaccine dose was 11 weeks (Range 2 to 12.7 weeks) and was not associated with Anti-S concentration after the second vaccine dose (p=0.22) (Supplementary figure 3b).

**Figure 3.**
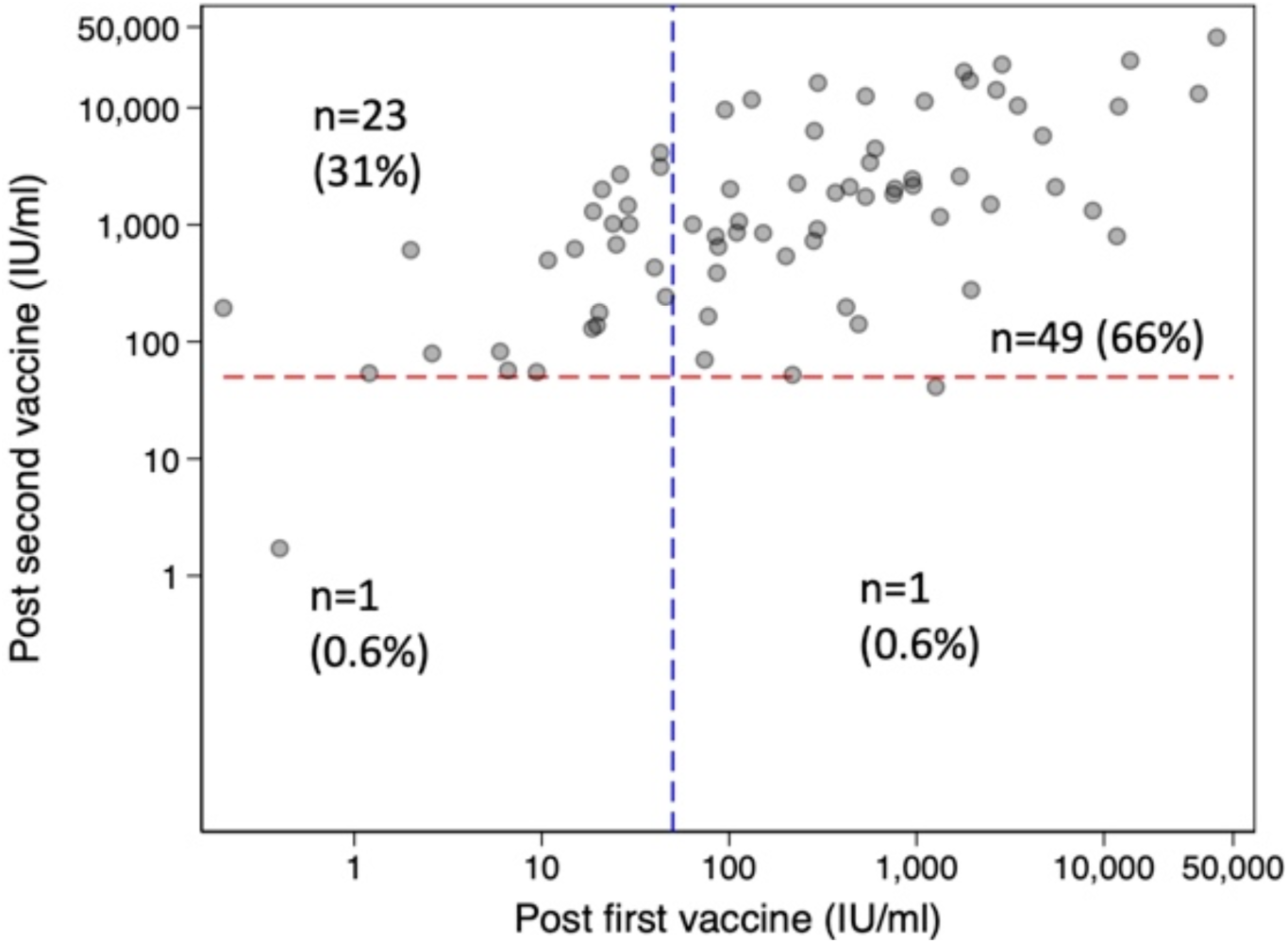
Relationship between Anti-S antibody concentration after 1st vs 2nd vaccine dose (n=74)

**Figure 4.**
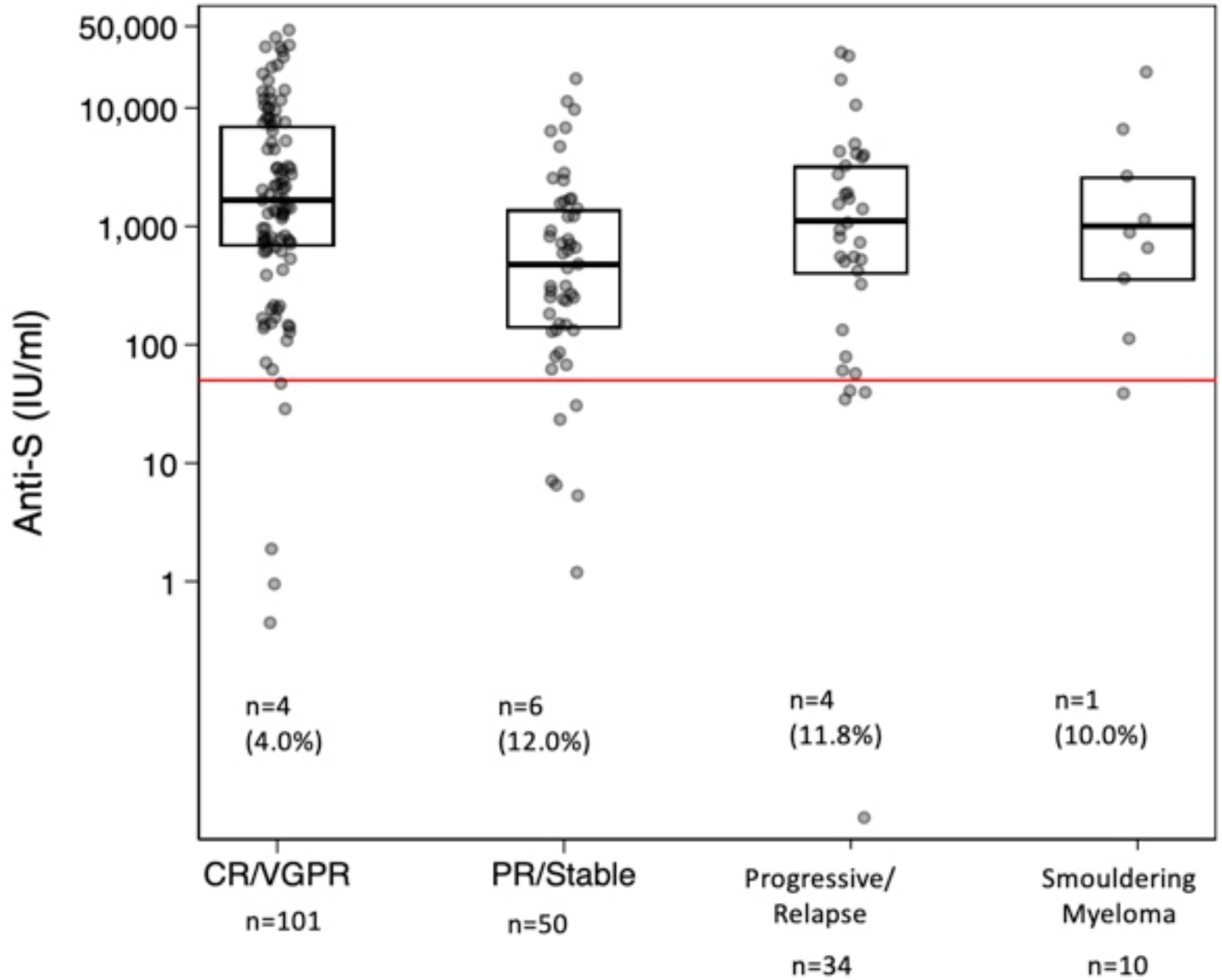
Relationship between Anti-S antibody concentration after 2nd vaccine dose and myeloma status from clinical records & patient reported (n=195) Legend: CR/VGPR – Complete remission/ Very good partial remission. PR/Stable – Partial remission/ Stable disease. Kruskal Wallis test, p<0.001 for myeloma patients only. For myeloma patients only (excluding smouldering myeloma patients)

Chemotherapy data was available for 126 patients at time of second vaccination. from the clinical records. Of the myeloma patients, 45 were on no treatment. Thirty-two patients were on CD38 antibody or BCMA containing regimens. Other non-CD38 regimens included proteasome inhibitors, immunomodulatory, alkylating chemotherapy and steroids (n=49). Those on chemotherapy had lower levels of Anti-S than those not on chemotherapy (p=0.025 (Figure 5). Using a threshold of 50 IU/ml to identify participants with suboptimal levels, the proportion of myeloma patients with < 50 IU/ml of Anti-S was non-significantly higher in those on chemotherapy (anti-CD38/ anti-BCMA – 3 and Other chemo - 5) vs those not on chemotherapy (8 (9.6%) vs 1 (2.2%), p=0.16).

**Figure 5.**
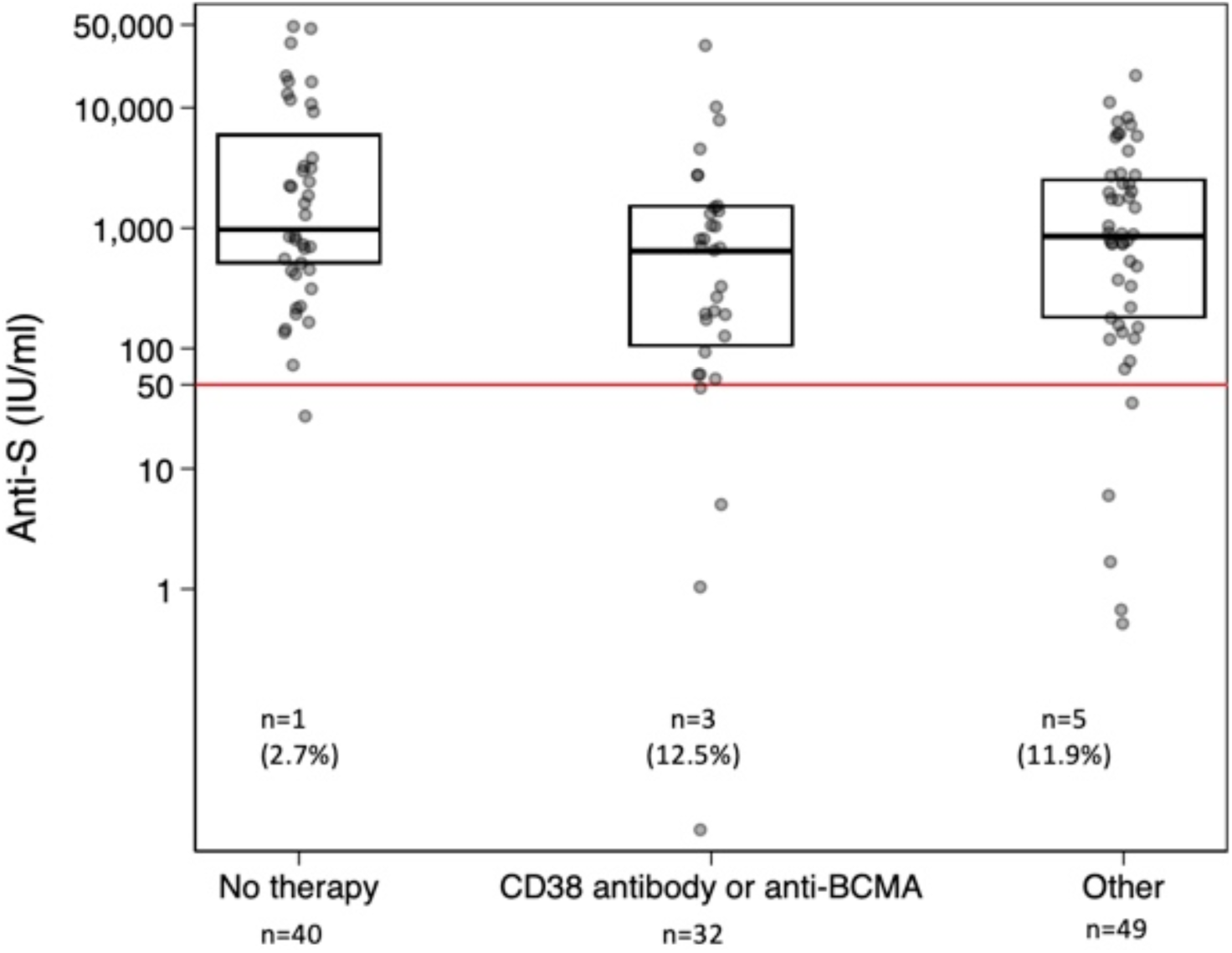
Relationship between Anti-S antibody concentration and chemotherapy at time of 2nd vaccine dose (n=121) Legend: CD38 antibody – daratumumab, Isatuximab. BCMA ADC-belantamab. Other: proteasome inhibitors - ixazomib, carfilzomib, bortezomib; immunomodulatory drugs - thalidomide, lenalidomide, pomalidomide; bendamustine, cyclophosphamide, dexamethasone, other steroids. bendamustine, cyclophosphamide, dexamethasone, other steroids. Kruskal Wallis for Anti-S level, p= 0.027. Fisher exact test for Anti-S <> 50 IU/ml, p=0.26

COVID-19 IGRA was measured after the second dose of vaccine, with results in 167/214 participants (Table 3). Positive IGRA results were significantly (p=0.002) associated with Anti-S serology status after second vaccine. Participants were more likely to be IGRA negative if they were not in CR/VGPR (p=0.021) with no significant differences by chemotherapy status (Table 4). Participants receiving the AZ vaccine had a significantly higher IGRA reactivity rate, 70.6% than those who received the PB 44.2% in all participants, and those with myeloma. (Table 5).

**Table 3.**
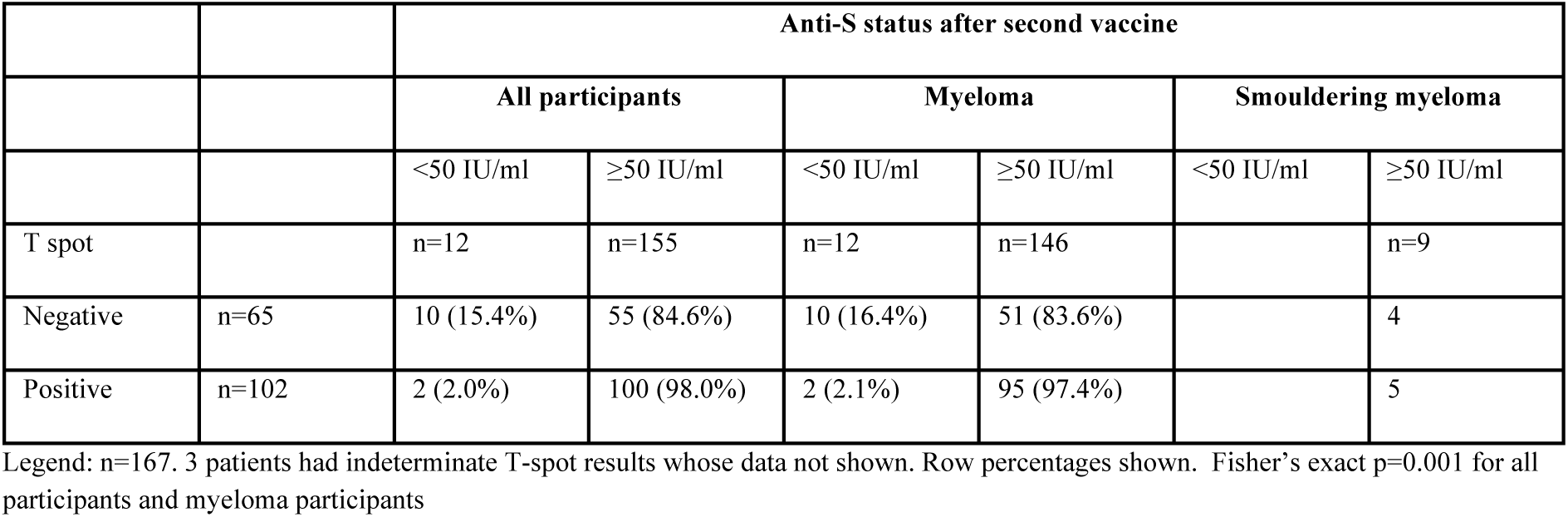
Relationship between S-Protein antibody status and T-cell response in all participants after the second COVID-19 vaccine dose.

|  |  | Anti-S status after second vaccine |  |  |  |  |  |
| --- | --- | --- | --- | --- | --- | --- | --- |
|  |  | All participants |  | Myeloma |  | Smouldering myeloma |  |
|  |  | <50 IU/ml | ≥50 IU/ml | <50 IU/ml | ≥50 IU/ml | <50 IU/ml | ≥50 IU/ml |
| T spot |  | n=12 | n=155 | n=12 | n=146 |  | n=9 |
| Negative | n=65 | 10 (15.4%) | 55 (84.6%) | 10 (16.4%) | 51 (83.6%) |  | 4 |
| Positive | n=102 | 2 (2.0%) | 100 (98.0%) | 2 (2.1%) | 95 (97.4%) |  | 5 |

Legend: n=167. 3 patients had indeterminate T-spot results whose data not shown. Row percentages shown. Fisher's exact p=0.001 for all participants and myeloma participants

**Table 4.** Relationship between IGRA reactivity and Myeloma status and Chemotherapy.

|  |  | Myeloma status (p=0.003) |  |  |  | Chemotherapy type (p=0.24) |  |  |
| --- | --- | --- | --- | --- | --- | --- | --- | --- |
|  |  | CR/VGPR | PR/Stable | Progressive<br>/ Relapse |  | No therapy | CD38/BCM<br>A | Other |
| T spot |  | n=76 | n=42 | n=22 |  | n=34 | n=24 | n=36 |
| Negative | n=55 | 20 (26.3%) | 23 (54.8%) | 12 (54.6%) | n=37 | 12 (35.3%) | 13 (54.2%) | 12 (33.3%) |
| Positive | n=85 | 56 (73.7%) | 19 (45.2%) | 10 (45.5%) | n=57 | 22 (64.7%) | 11 (45.8%) | 24 (66.7%) |

Legend: CR/VGPR – Complete remission/ Very good partial remission. PR/Stable – Partial remission/ Stable disease. Other chemotherapy included proteasome inhibitors - ixazomib, carfilzomib, bortezomib; immunomodulatory drugs - thalidomide, lenalidomide, pomalidomide; belantamab, bendamustine, cyclophosphamide, dexamethasone, other steroids. bendamustine, cyclophosphamide, dexamethasone, other steroids

**Table 5.** Relationship between IGRA reactivity and type of COVID-19 vaccine.

|  |  | All participants (n=119, p=0.002*) |  | Myeloma patients (n=111, p=0.006*) |  | Smouldering myeloma (n=8) (p=0.20**) |  |
| --- | --- | --- | --- | --- | --- | --- | --- |
| T spot |  | Oxford/AstraZeneca | Pfizer/BioNtech | Oxford/AstraZeneca | Pfizer/BioNtech | Oxford/AstraZeneca | Pfizer/BioNtech |
| Negative |  | 20 (28.2%) | 27 (56.3%) | 20 (29.4%) | 24 (55.8%) | 0 (0%) | 3 (60%) |
| Positive |  | 51 (71.8%) | 21 (43.8%) | 48 (70.6%) | 19 (44.2%) | 3 (100%) | 2 (40%) |

Legend: \*Chi-squared test. \*\*Fisher exact test

The multivariate models for determinants of an Anti-S concentration of less than 50 IU/ml are shown in Table 6a. Women were significantly less likely to have a low Anti-S concentration (< 50 IU/ml) than men, even after adjusting for myeloma status. For IGRA reactivity, increased age but not gender predicted a negative result, as did CD38 antibody/ anti-BCMA Ab exposure and receiving PB vaccine (table 6b). The IGRA reactivity rate in participants who received the AZ vaccine remained significant after adjusting for age, sex and myeloma status (p=0.006) and borderline significant after adjusting for chemotherapy (p=0.051). When analysing the different combinations of Anti-S and IGRA status, progressive disease/ relapse myeloma status predicted double negative status (Relative risk ratio 9.6 (95% CI 1.44 – 63.07)) after adjusting for age and sex. Having a positive Anti-S level with a negative IGRA release assay was less common (p=0.004) with the AZ than PB vaccine (RRR 0.23 (0.09 – 0.63) after adjusting for age, sex and myeloma status.

**Table 6a.** Independent predictors of low Anti-S concentration (≤50 IU/ml) post second vaccination in Myeloma patients only.

| Predictor | Model 1 (n=203) |  | Model 2 (n=186) |  | Model 3 (n=120) |  |
| --- | --- | --- | --- | --- | --- | --- |
|  | Odds Ratio | 95% CI | Odds Ratio | 95% CI | Odds Ratio | 95% CI |
| Age (year) | 1.04 | 0.97- 1.12 | 1.01 | 0.95- 1.09 | 1.06 | 0.96 – 1.18 |
| Female Sex | **0.21** | **0.05- -0.96** | **0.21** | **0.04- 1.00** | 0.22 | 0.039– 1.29 |
| Time between vaccine doses (weeks) | 0.8 | 0.63- 1.13 |  |  |  |  |
| Time from second vaccine dose and sample (weeks) | 0.92 | 0.78 – 1.08 |  |  |  |  |
| Myeloma status -partial response/ stable<sup>1</sup> |  |  | 3.37 | 0.86- 13.24 | 2.37 | 0.50 – 11.19 |
| Myeloma status - progression/ relapse<sup>1</sup> |  |  | 2.81 | 0.62 – 12.76 | 3.27 | 0.26 – 40.55 |
| CD38/ BCMA<sup>2</sup> |  |  |  |  | 5.03 | 0.63- 56.42 |
| Other chemo<sup>2</sup> |  |  |  |  | 5.98 | 0.63 – 56.42 |
| Pseudo-R<sup>2</sup> | 0.09 |  | 0.10 |  | 0.15 |  |

Legend: <sup>1</sup>Complete Remission/Very good partial response is comparator; <sup>2</sup>No chemotherapy is comparator; <sup>3</sup>PfizerBioNTech is comparator

**Table 6b.** Independent predictors of Negative T-spot post second vaccination in Myeloma patients only.

| Predictor | Model 1 (n=157) |  | Model 2 (n=105) |  | Model 3 (n=68) |  |
| --- | --- | --- | --- | --- | --- | --- |
|  | Odds Ratio | 95% CI | Odds Ratio | 95% CI | Odds Ratio | 95% CI |
| Age (year) | **1.04** | **1.01- 1.09** | 1.03 | 0.98 1.09 | 1.04 | 0.97- 1.11 |
| Female Sex | 0.84 | 0.43- 1.64 | 1.05 | 0.43- 2.60 | 1.06 | 0.32- 3.77 |
| Time between vaccine doses (weeks) | 1.02 | 0.85 – 1.64 |  |  |  |  |
| Time from second vaccine dose and sample (weeks) | 0.98 | 0.89-1.07 |  |  |  |  |
| Oxford/ AstraZeneca vaccine<sup>3</sup> |  |  | **0.27** | **0.11- 0.69** | 0.29 | 0.08 – 1.01 |
| Myeloma status - partial response/ stable<sup>1</sup> |  |  | **2.88** | **1.04 – 7.97** | 2.41 | 0.67- 8.73 |
| Myeloma status - progression/ relapse<sup>1</sup> |  |  | 2.30 | 0.76- 6.96 | 0.93 | 0.13 – 6.52 |
| CD38/ BCMA<sup>2</sup> |  |  |  |  | **4.80** | **1.09 -21.02** |
| Other chemo<sup>2</sup> |  |  |  |  | 1.21 | 0.31 – 4.70 |
| Pseudo-R<sup>2</sup> | 0.08 |  | 0.12 |  | 0.15 |  |

Legend: <sup>1</sup>Complete Remission/Very good partial response is comparator; <sup>2</sup>No chemotherapy is comparator; <sup>3</sup>PfizerBioNTech is comparator

## Discussion

Myeloma patients have been shielding or self-isolating since the start of the pandemic. Although the vast majority of the western population is now vaccinated, response to vaccination has been variable and myeloma patients require robust response to change their behaviour. This study has shown that following 2 doses of COVID-19 vaccination, myeloma patients can elicit anti-S protein antibodies in a significant proportion (92.7%) of patients. Our study despite its virtual consenting platform and requirement of IT literacy has recruited 32.7% of patients aged 70 and over, with representative ethnicity, therefore results can be extrapolated to the wider UK population. T cell responses also contribute to immunity and only a proportion of patients have elicited T cell responses after 2 doses. T cell responses following vaccination are not routinely studied and it is less clear how this would affect ability of patients to neutralise SAR-CoV2 infection. But a very small proportion of myeloma patients, 10/158 (6.3%) lacked both humoral and T cell response to vaccination. These patients should be directed to salvage strategies such as further doses of vaccination and passive antibody therapy to prevent severe COVID-19 (Supplementary figure 4). Patients showed a significant uplift of antibody response to second vaccination dose in 31% of patients (Supplementary figure 1). There are ongoing trials of passive antibody treatment for patients with poor antibody response ^12^.

Patients with smouldering myeloma elicited a robust anti-S antibody response to one dose of the vaccine, with one patient losing antibody response following Rituximab therapy for concomitant arthritis. This highlights that the durability of immune response can be affected by immunosuppressive agents. Prior COVID-19 infection significantly improved response to COVID-19 vaccination which has been observed in other haematological oncology patients^8^. One concern was the ability of AZ vaccine which is viral vector based in eliciting immune responses in myeloma patients. We found 2 doses of AZ vaccine gave a similar high proportion of patients a satisfactory humoral response using a threshold of >50 IU/ml as the Pfizer BioNtech vaccine, but with a higher T cell response in myeloma patients. Previously following one dose of PB vs AZ vaccination in 165 elderly immunosenescent population (80 year or older) significantly better T cell responses were noted after one dose of AZ vaccination. Our study extends this observation to two doses. The clinical implication of this is currently unclear and whether AZ vaccination induces better durability of both humoral and T cell response requires further follow up data ^13^. Despite there being 12-week dosing interval imposed in the UK we were not able to demonstrate a significant difference in anti-S responses between patients with increasing intervals between the first and second vaccine dose. Samples were obtained at varying time points in patients following the second dose of vaccine, significant difference was seen in relation to time of sample post vaccine, with falling anti spike antibody response with time, suggesting decay This requires further follow up and could be mitigated by planned third primary dose vaccination 6 months post second dose for myeloma patients in the UK. We have planned further sampling to assess durability of immune response prior to 3^rd^ dose (Figure 1). Both T cell and anti-S antibody response were dampened in patients who had partial response/ stable disease or had relapsing / progressing disease. Poor disease control is a predictor of double negative (T cell and humoral) immune response. This suggests that robust disease control should be secured prior to vaccination, or vaccination prior to intensive treatment, particularly during future booster vaccination doses in patients. We found up to 12% of patients who were on therapy that had low titres below threshold of anti-S antibody in comparison with patients not on therapy, but this was not significant. There was a similar non-significant trend noted in T cell response data analysis.

Previous studies have shown robust antibody responses in comparable proportion of myeloma patients when measured on different antibody assay platforms to that reported here^8,9,10^. Data from Ehsam et al. show up to 50% of myeloma patients elicited a T cell response^10^. This showed a good correlation with anti-S antibody response as observed in our patient population. Data from Terpos et al. who reported on neutralisation antibody responses showed only approximately 70% of myeloma patients have detectable levels^14^. This may be a better functional assay to use but not available in routine practice. In a small cohort of myeloma patients Bitoun et al. show a good correlation between titres of Anti-Spike IgG levels and neutralisation ability. If this data is confirmed in larger cohorts and neutralisation against VOC, a widespread use of humoral anti spike antibody in routine practice can be considered^15^. Both Terpos et al. and Van Oeklen et al. have shown data that ongoing therapy particularly with anti CD38 antibodies and BCMA targeted therapy significantly reduced antibody responses in myeloma patients^9,14^. We observed a higher proportion of patients on therapy having lower antibody levels, but we could not find a difference between the types of antimyeloma therapies administered. There may be differences in how these therapies have been applied (combinations), doses and/ or duration of therapy which may explain the variability observed.

Data from Terpos et al and Greenberger et al confirm our observation of better humoral responses to vaccination in women compared to men. It is currently unclear why there is differential immune response observed. But immune response to natural SARS-CoV-2 infection is also variable with a dominant adaptive immune response observed in women compared to men^16^. In comparison with immune response data reported following vaccination in lymphoma and CLL patients, myeloma patients are able to elicit better humoral and T cells responses^8,10^. But a small proportion of patients (6%) are antibody and IGRA negative who require salvage strategies to

### Strengths and weaknesses of the study

The strength of the study is confirmation of an immune response in heterogeneously vaccinated patients with longer than recommended duration between first and second dose. Our humoral response results are comparable to data reported so far. We have generated a large dataset of T cell response alongside humoral response in myeloma patients. Our data has also been generated in a comparable demographic to the UK myeloma population. Studies use different platforms to generate both antibody and T cell results which makes direct comparability difficult. This dataset, although very reassuring, requires longitudinal follow up. Although further vaccine doses are planned for this patient population, attrition of immune response over time due to ongoing therapy or underlying disease is a concern. A major limitation of this study is the limited sample size of participants with poor humoral and T cell responses. Further, our study is limited by the missing data on chemotherapy and myeloma status from the clinical notes for all participants. We partially addressed this by adding self-reported myeloma status where clinical data were not required and when we compared using only clinical record data, the findings were similar.

Therefore, further follow up to determine whether immune response wanes over time and whether use of particular therapies have a more significant detrimental effect on vaccine response requires evaluation. A subset of patients who require alternative strategies to prevent COVID-19 such as passive antibody therapy 12 or prophylactic antiviral therapy should be studied in prospective trials in this patient population.^17^. Whist these laboratory results are reassuring, the clinical implication of the observed humoral/cellular immune responses and real-world infection rates and severity of infection requires remains unanswered and highlight the complexity of the immune responses to different COVID-19 vaccines.

In conclusion, a robust humoral (92.7) anti-S antibody response can be elicited following either AZ or Pfizer mRNA vaccination in myeloma patients. 12-week dosing interval is not detrimental to immune response. Encouragingly a good proportion (60.1%) have also elicited T cell response. AZ vaccination provides robust T cell responses in myeloma patients. Data on durability of immune responses and including role of factors such as ongoing therapy, further vaccine doses require further follow up. Ongoing collection of data during patient follow up, including incidence of SARS-CoV2 infection and severity of COVID-19 would provide further clinical significance of the immune response elicited by vaccination, for myeloma patients and their carers.

## Data Availability

All data produced in the present work are contained in the manuscript

## Contributors

All listed authors made substantial contributions to the conception or design of the work; or the acquisition, analysis, or interpretation of data for the work; and drafting the work or revising it critically for important intellectual content; final approval of the version to be published; and agreement to be accountable for all aspects of the work in ensuring that questions related to the accuracy or integrity of any part of the work are appropriately investigated and resolved. KR is the guarantor and accepts full responsibility for the work and/or the conduct of the study, had access to the data, and controlled the decision to publish. The lead authors (KR, RS MKJ) designed the study and wrote the report. All authors were study investigators. SJ and PW were patient advisors. RS, SV, AT, JL and OC collated the data. KR, RS and MKJ analysed the data. All authors contributed to the review of the manuscript and approved the final version before submission.

## Declaration of Interests

All authors completed the ICMJE disclosure form. The following personal or financial relationships relevant to this manuscript existed during the conduct of the study. MD reports shares in Abingdon Health. SEM reports honoraria from Takeda, Janssen, Sanofi and Celgene. GC reports advisory board from Takeda, Celgene, Janssen, Sanofi, Oncopeptide, Roche, Karyopharm, IQVIA and Amgen. GC also reports research grant from Celgene, Takeda and IQVIA. NG reports honoraria from Janssen and Amgen and research grant from Kyowa Kirin. SuB reports advisory board from Pfizer and SANOFI. KMJ reports research grant from Amgen. KR reports honoraria, research grant from Janssen, Celgene, Takeda and Amgen. He also reports advisory board from Celgene, Takeda, Janssen, Amgen, Abbvie, Sanofi, Oncopeptides, Karyopharm, GSK, Adaptive biotech, Pfizer and speaker’s bureau from Celgene, Takeda and Adaptive Biotech.

## Data Sharing

Patients have consented to anonymised data sharing for research and publication purposes.

## Acknowledgements

Blood cancer UK, Myeloma UK patient panel, RUDY patient panel. We would also like to thank, Dr Stephen Jenkins (Russells Hall Hospital Dudley), Dr Eleni Tholouli (Manchester Royal Infirmary), Dr Richard Soutar (West of Scotland Cancer Clinic), Dr Bhuvan Kishore (Heartlands Hospital, University Hospitals Birmingham), Dr Firas Al-Kaisi (University Hospitals of Derby and Burton), Dr Paul Cervi (Southend University Hospital), Lucy Lunn (New Cross Hospital), Dr Mamta Garg and Laura Trotter (Leicester Royal Infirmary), Dr Jhansi Muddana and Juli Wilson (George Eliot Hospital), Dr Salim Shafeek (Worcestershire Acute Hospitals), Dr George Cherian (The Strewsbury and Telford Hospital), Dr Katie Randall and Dr Carolina Arbuthnot (Warwick Hospital), Dr Kamaraj Karunanithi (Royal Stoke University Hospital), Prof Guy Pratt (Queen Elizabeth Hospital) and Dr Hannah Hunter (Derriford Hospital) for their valuable contribution.

## Public and patient involvement statement

RUDY patients forum, Oxford Blood Group, Myeloma UK patient research panel were all consulted, and feedback secured on study design, questionnaire.

## Ethics approval

The study is based on the existing RUDYstudy.org platform (LREC 14/SC/0126 & RUDY LREC 17/SC/0501), an established online rare disease platform with online dynamic consent and patient reported outcome assessments. IRAS no: 213780, RUDY study Minor amendment 3, HRA approval 28th January 2021.

## Transparency declaration

Karthik Ramasamy, the lead author (and the manuscript’s guarantor), affirms that the manuscript is a honest, accurate, and transparent account of the study being reported; that no important aspects of the study have been omitted; and that any discrepancies from the study as planned (and, if relevant, registered) have been explained.

## Supplementary Figures

**Supplementary Figure 1.**
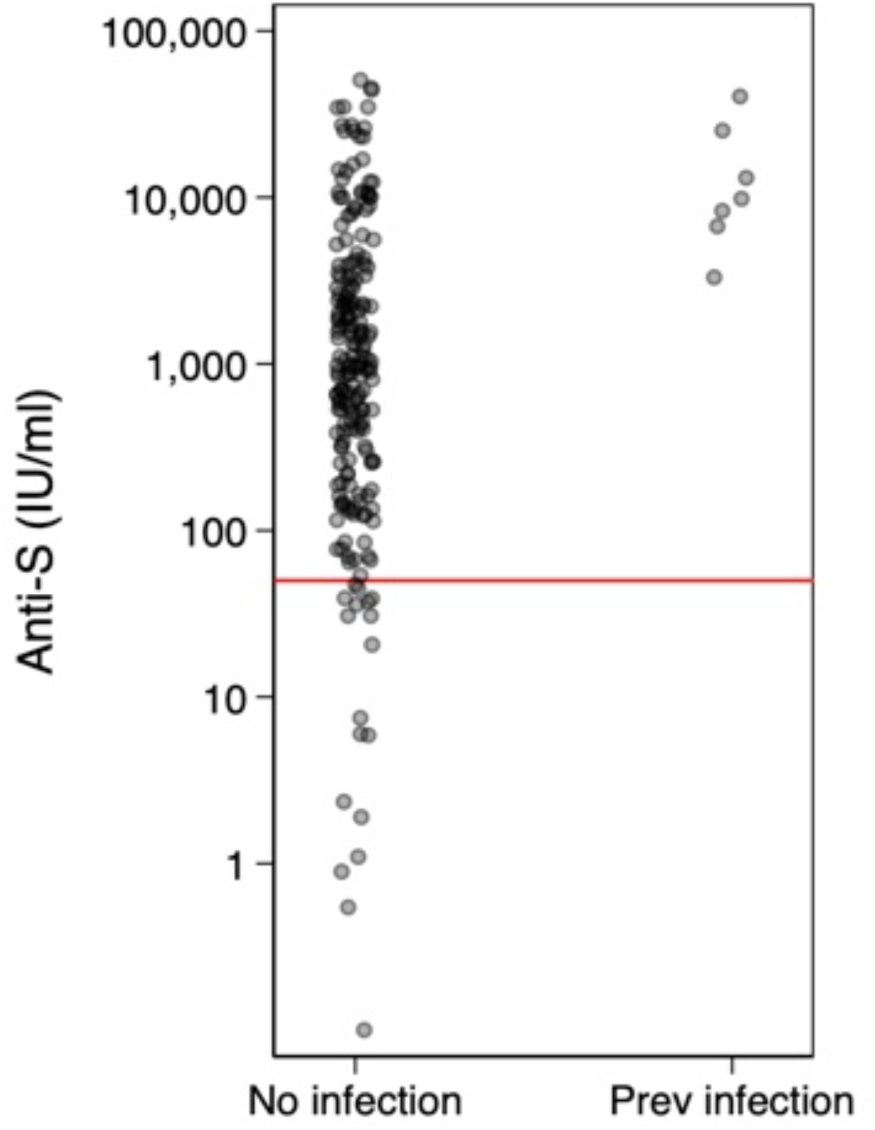
Relationship between Anti-S concentration after second vaccine dose and serological evidence of past COVID infection. Legend: Kruskal Wallis test p<0.001l

**Supplementary Figure 2a.**
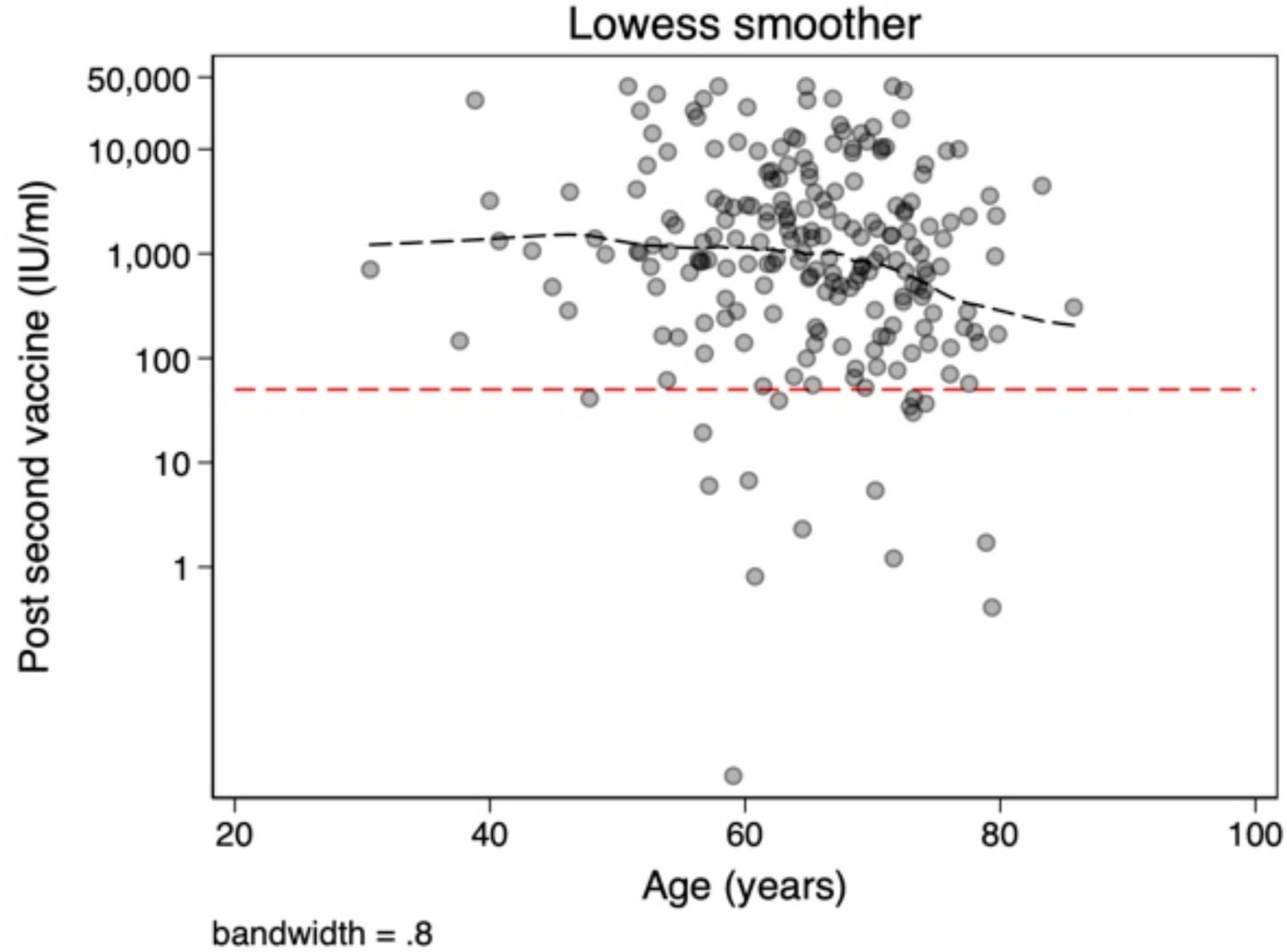
Relationship between Anti-S concentration after second vaccine dose and age. Legend: Lowess line shown. Spearman rho -0.15, p=0.021

**Supplementary Figure 2b.**
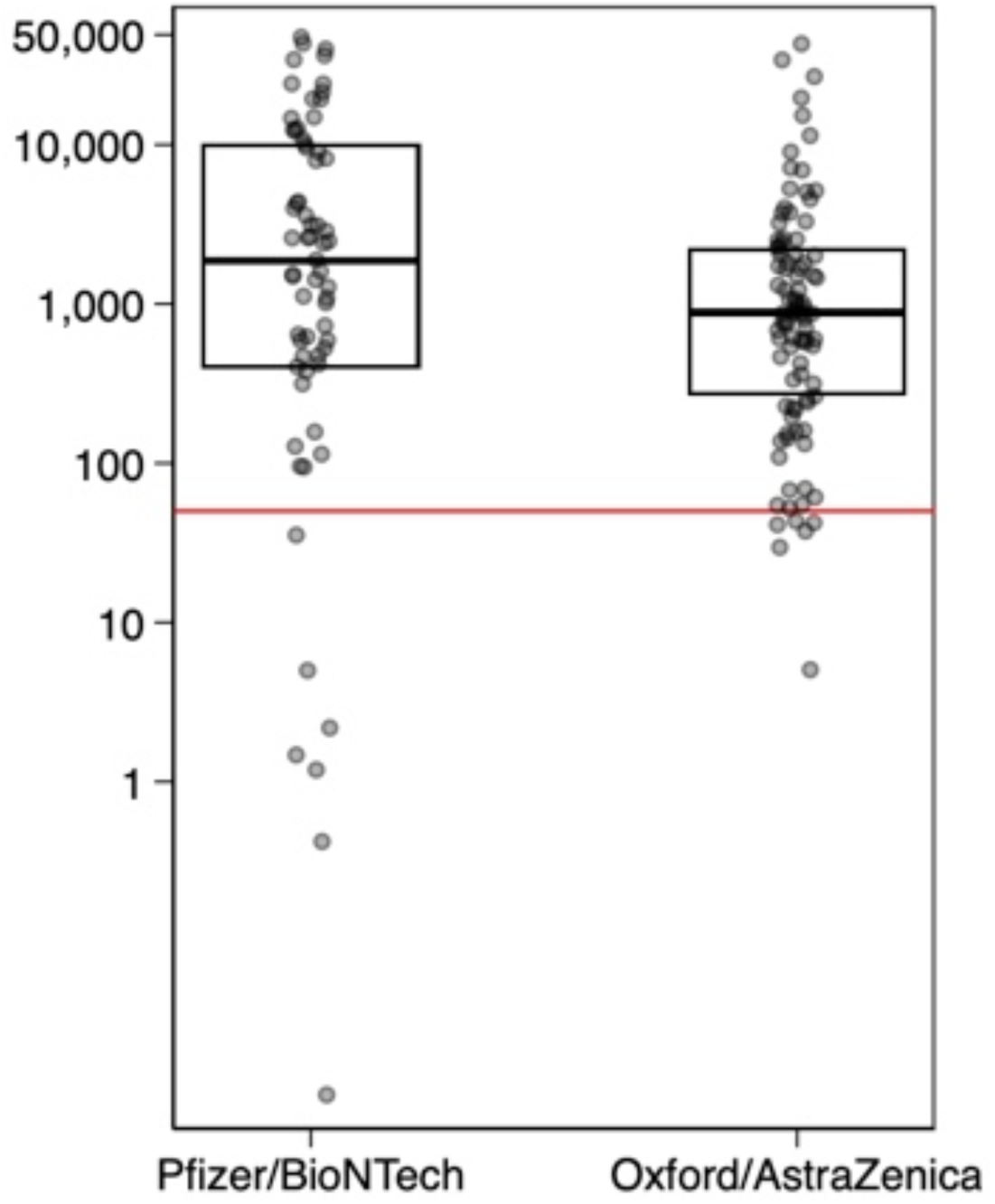
Comparison of Anti-S antibody after second vaccine dose by type of Vaccine. Legend: Comparison of Anti-S values < 50 IU/ml, using Fischer exact test, p=0.39 Kruskal-Wallis p=0.018

**Supplementary figure 3a and 3b.**
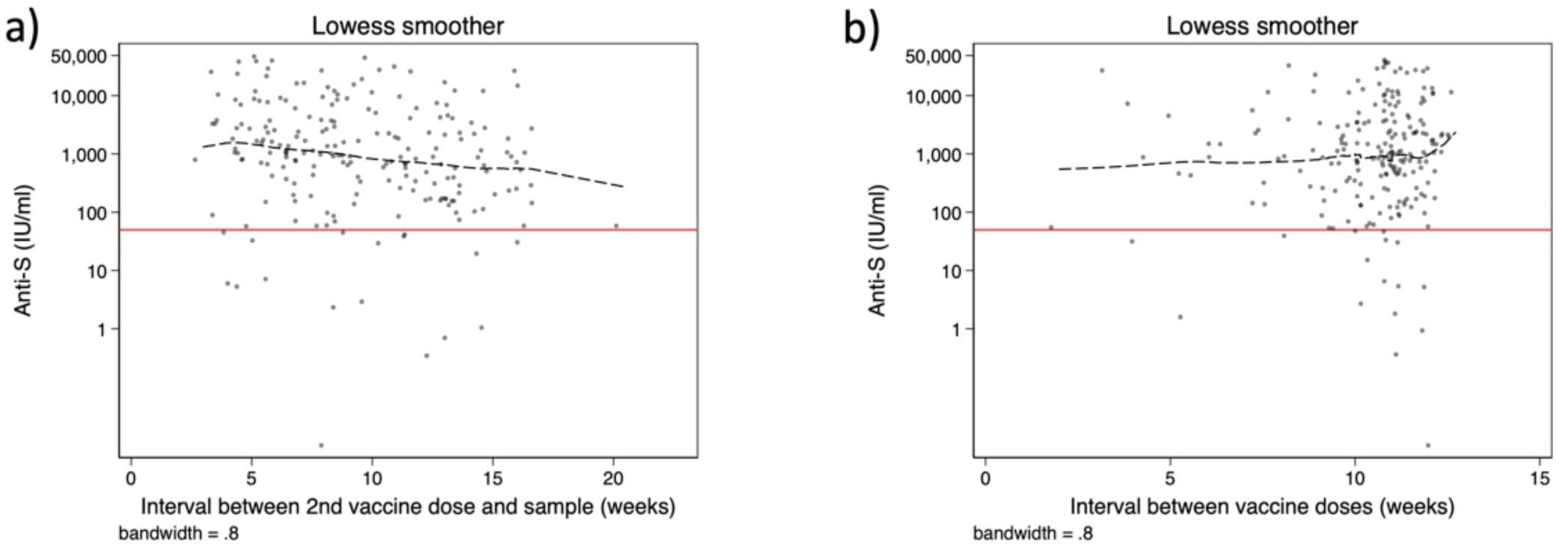
Relationship between Anti-S concentration and a) time from second vaccine; b) interval between vaccine doses. Legend: (a) Dash-lines represents Lowess line with 0.8 bandwidth, Solid red line, threshold of 50 IU/ml. Spearman’s rho= -0.12, p=0.002. (b) Dash-lines represents Lowess line with 0.8 bandwidth, Solid red line, threshold of 50 IU/ml. Spearman’s rho= 0.085, p=0.22

**Supplementary Figure 4.**
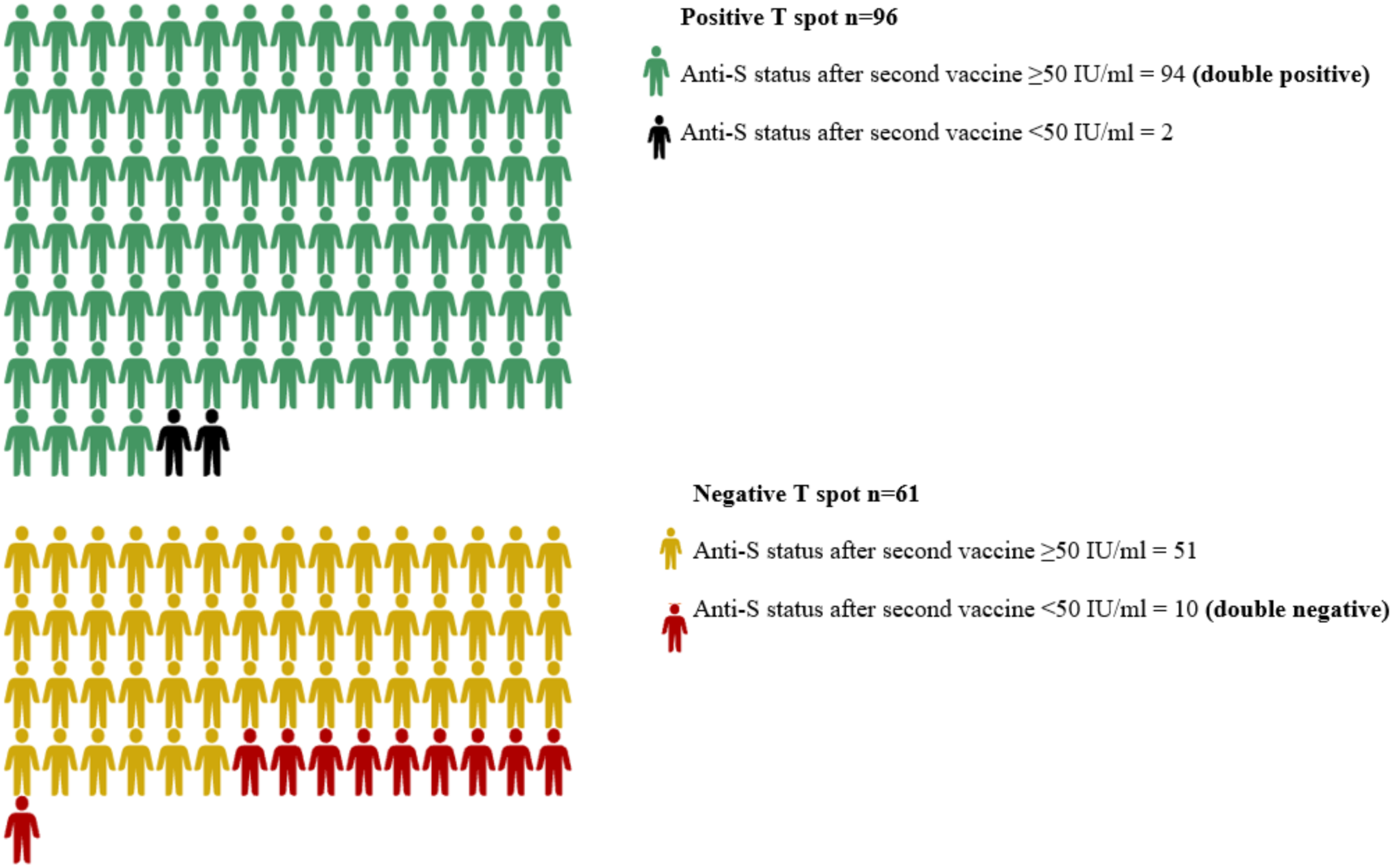
Visual representation of T-spot and humoral responses post vaccination.

## Notes

### Competing Interest Statement

The authors have declared no competing interest.

### Author Declarations

Ethics approval: The study is based on the existing RUDYstudy.org platform (LREC 14/SC/0126 & RUDY LREC 17/SC/0501), an established online rare disease platform with online dynamic consent and patient reported outcome assessments. IRAS no: 213780, RUDY study Minor amendment 3, HRA approval 28th January 2021 given by NHS HRA South Central - Berkshire B Research Ethics Committee.

